# Readmission Rates and Medication Selection for Patients with Heart Failure Preserved Ejection Fraction

**DOI:** 10.1101/2020.07.29.20163956

**Authors:** Troy Kramer, Carrie Vogler, Robert Robinson, Mukul Bhattarai

## Abstract

**Purpose:** Heart failure with preserved ejection fraction (HFpEF) has less guideline driven treatment options due to a lack of trials demonstrating medications with improved clinical outcomes for this patient population. The primary objective of this study is to determine which medications and dosages are related to high readmission rates for HFpEF patients.

**Methods:** A retrospective, single center, chart review was performed on patients with HFpEF at an academic medical center. Heart failure patients ages between 18-89 with an ejection fraction ≥45% from a transthoracic echocardiogram (TTE) were included. Primary outcomes include 30-day all cause readmission rates, prescribing patterns, and avoidance of potentially harmful medications. Descriptive statistics and multivariate logistic regression were used to assess potential risk factors.

**Results:** This study analyzed 455 patient admissions. Univariate analysis shows patients who were not readmitted were more likely to be on furosemide (54% vs 42%; p = 0.019). Conversely, readmitted patients were more likely to be taking bumetanide (4% vs 1%; p = 0.039). Lisinopril was the only angiotensin converting enzyme (ACE) inhibitor or angiotensin receptor blocker (ARB) associated with lower readmission rates (p = 0.036). Multivariate logistic regression showed bumetanide on admission (OR 14.6, p = 0.001), discharged on rosuvastatin (OR 6.29, p = 0.003) and meloxicam therapy (OR 6.33, p = 0.003) to be independent predictors of hospital readmission.

**Conclusion:** Three independent pharmacologic predictors for 30-day readmissions for patients with HFpEF were therapy with bumetanide, meloxicam, or rosuvastatin. Further research is needed to clarify the significance of these results.

## Introduction

### Background

Heart failure is a major public health concern in developed nations. In the United States, it is estimated that 5.7 million adults are living with some form of heart failure with an estimated total cost of 30.7 billion each year.^1,2^ There are two major categories of heart failure; heart failure with reduced ejection fraction (HFrEF) and heart failure with preserved ejection fraction (HFpEF). While both conditions include significant patient morbidity and mortality, treatment options are not as well defined for HFpEF. HFpEF is commonly referred to as diastolic heart failure and includes patients with heart failure symptoms with an ejection fraction ≥45%, though the exact ejection fraction percentage has been debated. HFpEF accounts for nearly 50% of heart failure patients.^3^ There are a limited number of trials that show medications with clinical improvement for HFpEF patients and few treatments recommended in current practice guidelines.^4,5,6^ Currently, the 2017 update to the AACF/AHA clinical practice guidelines recommends controlling blood pressure; however, it only gives moderate recommendations regarding agent selection with limited supporting data.^6^ The guidelines also recommend using diuretics for symptom control in volume overloaded patients.^6^ Further research is warranted to identify which medications result in the best clinical outcomes for patients and fewest number of hospitalizations.

HFpEF is a commonly encountered condition in the hospital setting, but the benefit of the medications prescribed and their impact on preventing hospital readmissions is not well studied. We studied the prescribing patterns of for HFpEF at Memorial Medical Center, a teaching hospital in Springfield, Illinois. An increase in knowledge regarding patient outcomes for various drug regimens will help to improve medication selection and reduce complications for future patients.

## Methods

### Study Design

This study was a single centered retrospective review of medications prescribed to patients with HFpEF. The study protocol was approved by the institutional review board prior to beginning the study. Patients reviewed included admission or discharge diagnosis ICD-9 or ICD-10 codes for diastolic heart failure (HFpEF) between 12/6/2016 and 12/6/2018. Data collection included recording admission and discharge medications that relate to heart failure for each patient. A comprehensive list of medications recorded can be seen in Tables 2-4. Medications that are commonly used for HFrEF were investigated for their potential use in patients with HFpEF. These medications were sourced from AACF/AHA 2017 clinical practice guidelines^6^. The chosen medications to avoid in heart failure examined in this trial are a subset of the 2016 AHA list of medications that may cause or exacerbate heart failure.^7^

**Table 1.** Baseline characteristics of the study population by 30-day readmission status.

|  | **Not readmitted within**<br>**30-days**<br>**n = 317** | **Readmitted**<br>**within 30-days**<br>**n = 128** | **p value** |
| --- | --- | --- | --- |
| **Age, mean (SD)** | 69 (12) | 67 (12) | 0.250 |
| **Female (%)** | 184 | 86 | 0.393 |
| **Length of stay (SD)** | 6.2 (5.12) | 6.16 (4.22) | 0.938 |
| **Hospital admissions in the last year**<br>**(SD)** | 0.24 (0.68) | 0.58 (1.42) | 0.008 |
| **Emergency department visits in last 6**<br>**months (SD)** | 0.13 (0.49) | 0.14 (0.43) | 0.805 |
| **Echocardiographic data** |  |  |  |
| **LVEF (SD)** | 57% (9.7) | 58% (9.9) | 0.185 |
| **Pulmonary Artery Pressure** | 44 (17.7) | 42 (15.2) | 0.399 |
| **E/e' Ratio** | 16.33 (8.2) | 15.14 (7.7) | 0.223 |
| **HOSPITAL Score (SD)** | 4.10 (1.38) | 4.73 (1.60) | <0.001 |
| **LACE Index (SD)** | 10.61 (1.87) | 10.80 (1.94) | 0.338 |
| **Charlson Comorbidity Score (SD)** | 3.39 (1.18) | 3.31 (1.10) | 0.489 |
| **Medical Comorbidities (%)** |  |  |  |
| **Myocardial infarction** | 71 | 23 | 0.165 |
| **Peripheral artery disease** | 30 | 8 | 0.194 |
| **Stroke** | 27 | 9 | 0.468 |
| **Dementia** | 10 | 3 | 0.564 |
| **Chronic lung disease** | 193 | 92 | 0.241 |
| **Connective tissue disease** | 14 | 5 | 0.697 |
| **Peptic ulcer disease** | 5 | 4 | 0.352 |
| **Cirrhosis** | 31 | 17 | 0.418 |
| **Diabetes without complications** | 130 | 51 | 0.417 |
| **Diabetes with complications** | 129 | 54 | 0.755 |
| **Paralysis** | 10 | 0 | 0.035 |
| **Renal disease** | 148 | 66 | 0.823 |
| **Cancer** | 20 | 13 | 0.240 |
| **Metastatic cancer** | 4 | 6 | 0.039 |
| **Admission laboratory values** |  |  |  |
| **Hemoglobin units (SD)** | 10.92 (2.47) | 10.57 (2.12) | 0.136 |
| **Platelet (units) (SD)** | 231 (88.7) | 251 (97.5) | 0.036 |
| **White blood cell count (SD)** | 10.53 (5.30) | 10.37 (5.01) | 0.757 |
| **Blood urea nitrogen (SD)** | 29.21 (19.3) | 30.30 (20.27) | 0.590 |
| **Creatinine (SD)** | 1.76 (1.39) | 2.11 (2.10) | 0.079 |
| **Potassium (SD)** | 4.21 (0.68) | 4.32 (0.760) | 0.153 |
| **Sodium (SD)** | 136.43 (4.86) | 135.80 (4.31) | 0.172 |
| **Body mass index (SD)** | 37.42 (15.49) | 35.72 (12.56) | 0.223 |

**Table 2.** Admission Medications by 30-day readmission status.

| Medication Class | Medication | Not readmitted within 30-days<br>N = 317 | Readmitted within 30-days<br>N = 138 | p value |
| --- | --- | --- | --- | --- |
| **ACE Inhibitors** | Benazepril | 1 (1%) | 1 (1%) | 0.544 |
|  | Enalapril | 2 (1%) | 0 | 0.350 |
|  | Lisinopril | 102 (32%) | 31 (22%) | **0.036** |
|  | Quinapril | 1 (1%) | 0 | 0.509 |
| **Angiotensin receptor blocker** | Losartan | 28 (9%) | 11 (8%) | 0.763 |
|  | Valsartan | 4 (1%) | 1 (1%) | 0.613 |
| **Beta Blockers** | Atenolol | 6 (2%) | 1 (1%) | 0.352 |
|  | Bisoprolol | 8 (3%) | 0 | 0.60 |
|  | Carvedilol | 63 (20%) | 27 (20%) | 0.939 |
|  | Labetalol | 1 (1%) | 2 (1%) | 0.170 |
|  | Metoprolol Succinate | 75 (24%) | 28 (20%) | 0.430 |
|  | Metoprolol Tartrate | 57 (18%) | 19 (14%) | 0.268 |
| **Cardiac Glycosides** | Digoxin | 12 (4%) | 6 (4%) | 0.77 |
| **Loop diuretics** | Bumetanide | 2 (1%) | 12 (9%) | **<0.001** |
|  | Furosemide | 172 (54%) | 52 (38%) | **0.001** |
|  | Torsemide | 26 (8%) | 5 (4%) | 0.075 |
| **Nitrates** | Isosorbide Dinitrate | 3 (1%) | 0 | 0.252 |
|  | Isosorbide Mononitrate | 34 (11%) | 18 (13%) | 0.475 |
| **Statins** | Atorvastatin | 111 (35%) | 46 (33%) | 0.729 |
|  | Lovastatin | 12 (4%) | 7 (5%) | 0.528 |
|  | Pitavastatin | 1 (1%) | 0 | 0.509 |
|  | Pravastatin | 32 (10%) | 13 (9%) | 0.825 |
|  | Rosuvastatin | 6 (2%) | 9 (7%) | **0.011** |
|  | Simvastatin | 19 (6%) | 14 (10%) | 0.117 |
| **Other** | HCTZ | 19 (6%) | 12 (9%) | 0.293 |
|  | Metolozone | 9 (3%) | 5 (4%) | 0.656 |
|  | Spironolactone | 31 (10%) | 12 (9%) | 0.716 |
|  | Triamterene | 4 (1%) | 4 (1%) | 0.22 |

### Patient Population

Eligible patients were between the ages of 18-89 with an admission to the internal medicine hospitalist service, and an ejection fraction of ≥ 45% reported on a TTE during admission or within previous 6 months. Patients who were on hospice, died during their hospital stay, had a past medical history of valvular heart disease, a history of cardiotoxicity due to chemotherapy, or receiving chemotherapy were excluded from the study.

### Outcomes

The primary efficacy outcomes were 30-day all cause readmission rates for patients with HFpEF. Other primary outcomes include selection of medications for HFpEF and avoidance of medications that should be avoided in heart failure patients. Secondary outcomes include patient safety measured by evaluating readmission rates between patients.

### Statistical analysis

A HOSPITAL score, LACE index, Charlson Comorbidity Score were calculated and compared for each admission. Qualitative variables were compared using Pearson chi^2^ or Fisher’s exact test and reported as frequency (%). Quantitative variables were compared using the non-parametric Mann–Whitney *U* test and reported as mean ± standard deviation. Variables from univariate analysis with a p value of 0.05 or less were evaluated using multivariate logistic regression with backwards likelihood ration based selection. Statistical analyses were performed using SPSS version 22 (SPSS Inc., Chicago, IL, USA). Two sided *P*-values < 0.05 were considered significant.

## Results

### Population Baseline

A total of 455 unique admissions were analyzed. Results for all cause readmissions within 30-days are listed in Tables 1-4. Baseline characteristics between those who were not readmitted within 30-days versus those who were readmitted within 30-days are summarized in Table 1. The two groups were comparable in their medical comorbidities as indicated by their Charlson Comorbidity index. Those who were readmitted within 30-days were more likely to have had hospital readmissions within the past year (p = 0.008). These patients were also more likely to have a higher HOSPITAL score (p < 0.001) with an odds ratio of 1.23 (1.05-1.44, 95% CI). The HOSPITAL score is a validated screening tool used to help identify potentially avoidable 30-day readmissions using seven defined attributes at patient discharge.^8^

### Medications on Admission

Four admission medications were found to be statistically significant in relation to 30-days all cause readmissions as summarized in Table 2. Lisinopril was the only angiotensin converting enzyme (ACE) inhibitor or angiotensin receptor blocker (ARB) associated with a lower rate of readmissions (p = 0.036). Two of the three medications in the loop diuretic class were found to have significant correlations with readmissions. The three loop diuretics that were investigated were furosemide, torsemide, and bumetanide. Patients coming to the hospital on furosemide were associated with a lower 30-day readmission (p = 0.001). Conversely, patients who were on bumetanide were associated with higher rates of 30-day readmissions (p < 0.001) with an odds ratio of 14.6 (3.12-68.67, 95% CI). There was no statistical difference for patients who were on torsemide. The final drug that was found to be statistically significant in correlation to 30-day all cause readmissions was rosuvastatin. Patients who were on rosuvastatin on admission were associated with a greater occurrence of 30-day readmissions (p = 0.011).

### Medications on Discharge

Discharge medications for patients with HFpEF and how they relate to 30-day all cause readmission rates is summarized in Table 3. Enalapril was associated with increased readmissions (p = 0.032) and was the only ACE inhibitor or ARB to show a statistically significant increase in readmissions for discharge medications. There were two loop diuretics that showed statistical significance with discharge medications for 30-day readmissions. Patients who were readmitted within 30-days were more likely to be taking bumetanide from their last discharge (p = 0.039) while patients readmitted within 30-days were less likely to be taking furosemide from their last discharge (p = 0.019). For other medications, rosuvastatin was the only other medication found to be statistically significant since patients readmitted within 30-days were more likely to be taking rosuvastatin than those who were not (p = 0.002) with an odds ratio of 6.29 (1.83-21.60, 95% CI).

**Table 3.** Discharge Medications by Readmission Status.

| Medication Class | Medication | Not readmitted within 30-days<br>N = 317 | Readmitted within 30-days<br>N = 138 | p value |
| --- | --- | --- | --- | --- |
| **ACE Inhibitors** | Benazepril | 2 (1%) | 0 | 0.350 |
|  | Enalapril | 0 | 2 (1%) | 0.032 |
|  | Lisinopril | 85 (27%) | 26 (19%) | 0.069 |
|  | Quinapril | 0 | 0 |  |
| **Angiotensin receptor blocker** | Losartan | 20 (6%) | 7 (5%) | 0.608 |
|  | Valsartan | 1 (1%) | 1 (1%) | 0.544 |
| **Beta Blockers** | Atenolol | 4 (1%) | 0 | 0.185 |
|  | Bisoprolol | 6 (2%) | 1 (1%) | 0.352 |
|  | Carvedilol | 83 (26%) | 30 (22%) | 0.313 |
|  | Labetalol | 1 (1%) | 3 (2%) | 0.051 |
|  | Metoprolol Succinate | 55 (17%) | 29 (34%) | 0.354 |
|  | Metoprolol Tartrate | 67 (21%) | 23 (17%) | 0.271 |
| **Cardiac Glycosides** | Digoxin | 15 (5%) | 6(4%) | 0.858 |
| **Loop diuretics** | Bumetanide | 4 (1%) | 6 (4%) | 0.039 |
|  | Furosemide | 171 (54%) | 58 (42%) | 0.019 |
|  | Torsemide | 24 (8%) | 5 (4%) | 0.113 |
| **Nitrates** | Isosorbide Dinitrate | 3 (1%) | 0 | 0.252 |
|  | Isosorbide Mononitrate | 28 (9%) | 17 (12%) | 0.252 |
| **Statins** | Atorvastatin | 126 (40%) | 52 (37%) | 0.678 |
|  | Lovastatin | 9 (3%) | 5 (4%) | 0.656 |
|  | Pitavastatin | 1 (1%) | 0 | 0.509 |
|  | Pravastatin | 26 (8%) | 13 (9%) | 0.670 |
|  | Rosuvastatin | 4 (1%) | 9 (7%) | 0.002 |
|  | Simvastatin | 17 (5%) | 12 (9%) | 0.181 |
| **Other Diuretics** | HCTZ | 13 (4%) | 5 (4%) | 0.810 |
|  | Metolozone | 6 (2%) | 0 | 0.104 |
|  | Spironolactone | 29 (6%) | 9 (7%) | 0.829 |
|  | Triamterene | 3 (1%) | 2 (1%) | 0.636 |

### Medications to Avoid in Heart Failure

Readmission rates for patients taking medications that should be avoided in heart failure are summarized in Table 4. Meloxicam was the only medication found to be associated with 30-day all cause readmissions for HFpEF patients (p = 0.002) with an odds ratio of 6.33 (1.87-21.43, 95% CI). Meloxicam may have this effect with its inhibition of prostaglandin synthesis promoting water retention.

**Table 4.**
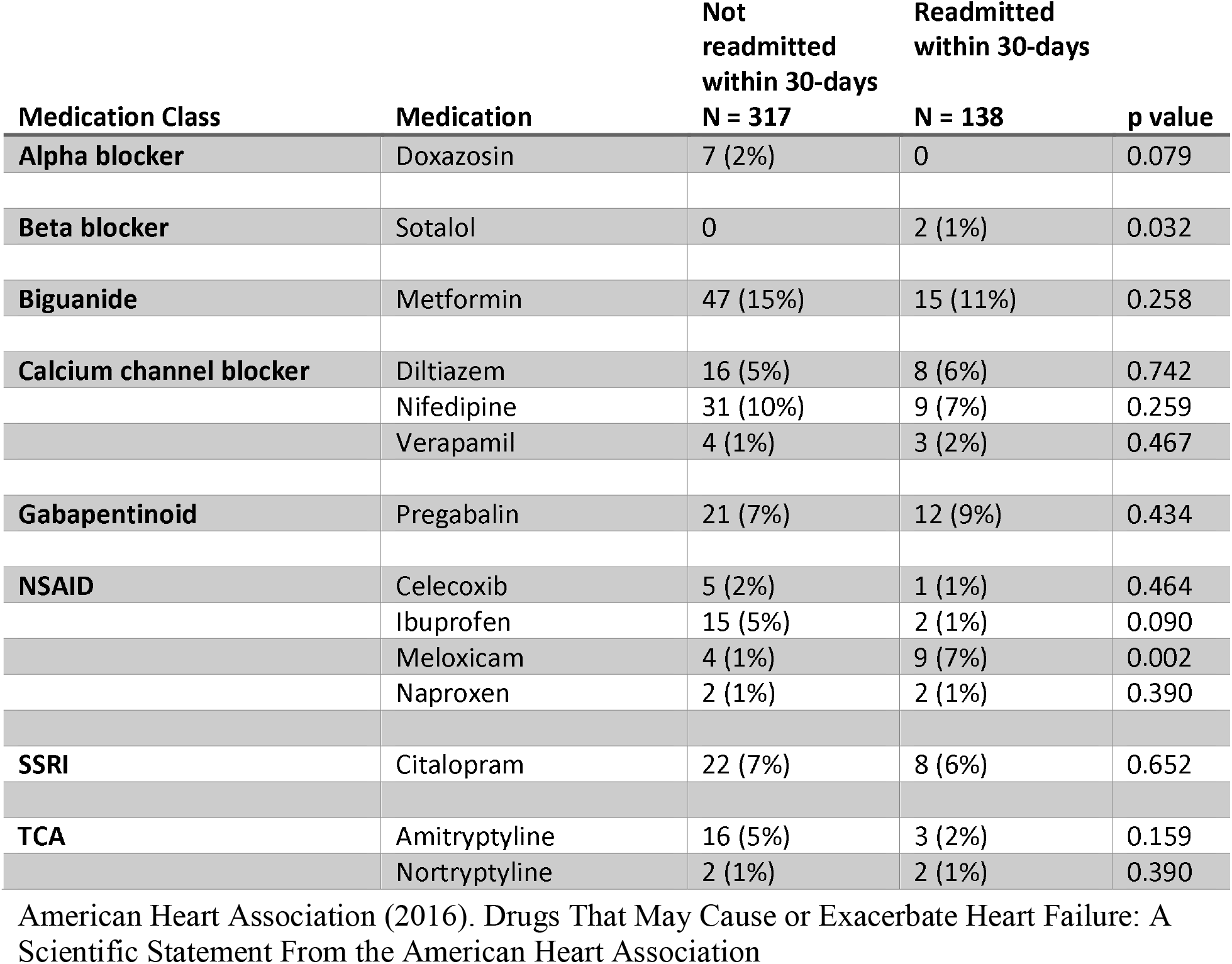
Medications to be avoided with HFpEF by Readmission Status.

### Multivariate analysis

Multivariate logistic regression showed bumetanide on admission (OR 14.6, p = 0.001), being discharged on rosuvastatin (OR 6.29, p = 0.003), meloxicam therapy (OR 6.33, p = 0.003), an elevated HOSPITAL score (OR 1.23, p = 0.011) and an elevated platelet count (OR 1.00, p = 0.011) to be independent predictors of hospital readmission.

## Discussion

Overall, lisinopril, enalapril, furosemide, bumetanide, rosuvastatin, and meloxicam had a statistically significant correlation with readmission rates for HFpEF patients. The difference in the loop diuretics appears to be the most clinically relevant finding. Furosemide was found to correlate with less 30-day readmission rates while bumetanide was found to correlate with more 30-day readmissions. A follow up comparing equivalent dosing between these two agents may be warranted. These findings with furosemide agree with the ACC/AHA/HFSA 2017 practice guidelines of using diuretics to symptomatic volume control for furosemide, but disagree with bumetanide because despite being an agent used for volume control, it was associated with higher readmissions.^6^

Rosuvastatin was the only member in its class to show significance in 30-day readmissions. While all statins were reviewed, one possible explanation is that the intensity of the statin could have an effect on patient outcomes. Rosuvastatin and atorvastatin both are indicated for both high intensity and moderate intensity statin therapy. Since atorvastatin did not show statistical significance in all cause 30-day readmissions, statin intensity may not be the reason for this result unless patients on rosuvastatin were more likely to be on a different intensity therapy than those taking atorvastatin. Analyzing readmission rates in relation to statin intensity would provide further insight into this.

Other drugs commonly used in patients with HFrEF including ACE inhibitors, ARBs, and beta blockers did not show any statistically significant correlation with 30-day readmissions. One previous study had shown the potential for candesartan to have a moderate impact in reducing hospitalizations in patients with HFpEF.^9^ While this study did not look directly at candesartan, there were two other ARBs examined. This study did not find any statistically significant correlation between patients on ARBs and 30-day readmissions.

For the use of aldosterone antagonists in HFpEF, it was found that there was no statistically significant correlation between their use and 30-day all cause readmissions. These results are consistent with the TOPCAT trial evaluating the use of spironolactone for HFpEF patients. In this prospective double-blind placebo-controlled trial, it was found that spironolactone did not significantly impact all cause hospitalization rates (18.8 hospitalizations per 100 person-years in the spironolactone group and 20.0 per 100 person-years in the placebo group, (hazard ratio 0.94 (0.85-1.04, 95% CI, p=0.25))^10^. The TOPCAT trial did, however, show significance for reducing hospitalizations for heart failure, (3.8 hospitalizations for heart failure per 100 person-years in the spironolactone group vs. 4.6 per 100 person-years in the placebo group; hazard ratio 0.83 (0.69 to 0.99, 95% CI, p=0.04)), which may mark an area for further study^10^.

One of the limitations to this study is the sample size. Many of the drugs analyzed had a relatively small number of patients taking them which increases the likelihood of a type 2 error. Another limitation involves safety outcomes with nonsteroidal anti-inflammatory drugs (NSAIDs). It is possible that patients may take over the counter NSAIDs without reporting them in their medication list which makes them difficult to track. Patients using these medications to help with pain relief may take them as needed which makes them difficult to assess for use. Medications adherence and side effects were not assessed in this study which could impact the readmission rates if patients were not talking their diuretics as prescribed. Strengths of the study are that it was a single centered and had one hospitalist group which reduces variability in documentation and charting of patient data.

## Conclusion

In the study there were four identified potential risk factors for 30-day readmissions for patients with HFpEF as shown in Table 5. These were found to be HOSPITAL score, patients on bumetanide on admission, discharge on rosuvastatin, and patients on meloxicam on admission.. In treatment of patients with HFpEF, furosemide appeared to be associated with the lowest frequency of readmissions. This suggests that prescribing patterns of specific loop diuretics and may make a significant impact in preventing patients from becoming readmitted.

**Table 5.** Multivariate logistic regression of potential risk factors for hospital readmission within 30-days of discharge.

|  | Odds Ratio (95% CI) | P value |
| --- | --- | --- |
| HOSPITAL score | 1.23 (1.05 – 1.44) | 0.011 |
| Bumetanide on admission | 14.6 (3.12 – 68.67) | 0.001 |
| Discharged on Rosuvastatin | 6.29 (1.83 – 21.60) | 0.003 |
| Meloxicam therapy | 6.33 (1.87 - 21.43) | 0.003 |
| Platelet count | 1.00 (1.00 – 1.01) | 0.011 |

## Data Availability

Data obtained during this study was accessed with IRB approval.

## Acknowledgements

Nicolas Ferry, SIU Medical Student for his assistance in data collection.

## Author contributions

All authors contributed to this work.

